# Characteristics of health-related quality of life and related factors in patients with brain tumors treated with rehabilitation therapy

**DOI:** 10.1101/2022.04.25.22274293

**Authors:** Takahiro Watanabe, Shinichi Noto, Manabu Natsumeda, Shinji Kimura, Satoshi Tabata, Fumie Ikarashi, Mayuko Takano, Yoshihiro Tsukamoto, Makoto Oishi

## Abstract

**Background:** Rehabilitation therapy during hospitalization is effective in improving activities of daily living (ADL) and physical function in patients with brain tumors. However, there are few studies on the effect of rehabilitation therapy on health-related quality of life (HRQOL) in patients with brain tumors. Additionally, the EuroQol-5Dimension-5Level (EQ-5D-5L) index score has not been reported as an outcome. This study aimed to investigate the HRQOL of patients with brain tumors who underwent rehabilitation therapy and investigated the factors affecting the EQ-5D-5L index score from various perspectives, including various brain tumor types, treatment methods, and recurrence. In addition, we examined the relationship between the EQ-5D-5L index score, disease-specific HRQOL scale, and ADL.

**Methods:** Patients with brain tumors who underwent treatment and rehabilitation at Niigata University Medical & Dental Hospital were included in this cross-sectional study. We used the EQ-5D-5L, European Organisation for Research and Treatment of Cancer (EORTC) quality of life questionnaire core 30, and EORTC quality of life questionnaire brain cancer module to evaluate HRQOL. ADL were assessed using the functional independence measure (FIM). The relationship between each HRQOL assessment score and the FIM was analyzed, and the influence of related factors was assessed by multiple regression analysis.

**Results:** This study included 76 patients. The EQ-5D-5L index score was 0.689 for all patients with brain tumors and 0.574 for those with glioblastomas, which was the lowest value. There was a strong correlation between the EQ-5D-5L index score and FIM (r = 0.627, p<0.001). In addition, the EQ-5D-5L index score was significantly correlated with most of the items of the disease-specific HRQOL scale. Multiple regression analysis revealed that glioblastoma histology (coefficient: -0.570, p = 0.024) and surgery (coefficient: 0.376, p = 0.030) were independent factors affecting the EQ-5D-5L index score.

**Conclusions:** Patients with glioblastoma undergoing rehabilitation may have reduced HRQOL, which was influenced by glioblastoma histology and surgery.

## Introduction

Brain tumors are broadly classified as primary and metastatic brain tumors (MBTs), the latter of which are most commonly caused by the metastasis of lung cancer or breast cancer. Primary brain tumors affect approximately 7 people per 100,000 population worldwide every year, and the incidence is on the rise [1]. The treatment of brain tumors varies depending on the tumor category, generally consisting of multidisciplinary treatment with surgery, radiation therapy, and chemotherapy [2]. There has been great progress in the treatment methods for brain tumors in recent years, which have prolonged the survival of patients with brain tumors. Nevertheless, in some malignant brain tumors, the prognosis remains poor even with the aforementioned treatments. In particular, glioblastoma recurs at 6 months to 1 year on average. The reported median overall survival (OS) in glioblastoma is >1.5 years, but the 5-year survival rate is still only 15% [2]. Similarly, the median OS for MBTs is 12.0 months, even for prostate cancer, which is considered to have the longest OS [3]. In patients with difficult-to-cure brain tumors and poor prognoses, it is important to improve and maintain health-related quality of life (HRQOL), which is a patient-reported outcome, as well as OS and progression-free survival.

The European Organisation for Research and Treatment of Cancer (EORTC) quality of life questionnaire core 30 (QLQ-C30) and EORTC quality of life questionnaire brain cancer module (BN20) are frequently used in the assessment of HRQOL in patients with brain tumors [4,5]. These HRQOL scales are intended to capture disease-specific psychosomatic functions and symptoms. In recent years, the need for the economic evaluation of medical treatments has been increasing worldwide, generating more interest in utility scales. The EuroQol-5Dimension-5Level (EQ-5D-5L) is one of the most popular scales for calculating the utility index [6], but only a few studies have employed this scale in the evaluation of patients with brain tumors [7-9].

Rehabilitation therapy during hospitalization is reportedly effective in improving activities of daily living (ADL) and physical function in patients with brain tumors. Studies comparing patients with brain tumors to those with stroke and cerebral infarction with similar symptoms found comparable improvements in physical function, ADL, and home discharge rates [10-13]. In addition, even in patients with glioblastomas and MBTs, who are considered to have poor prognoses, a significant improvement in their total functional independence measure (FIM) score has been reported after rehabilitation[14,15].

In contrast, there are few previous studies on the effect of rehabilitation therapy on HRQOL in patients with brain tumors [16]. Furthermore, the efficacy of rehabilitation therapy on HRQOL has not been examined in detail [17-20]. Additionally, the EQ-5D-5L index score has not been reported as an outcome. Thus, the efficacy of rehabilitation therapy differs between patients with brain tumors and those with stroke in terms of ADL and HRQOL, but the relationship between the two has not been fully investigated. In addition to the impact of disease factors such as brain tumor type and recurrence, few previous studies have analyzed the multifaceted impact of treatments such as surgery, radiotherapy, and chemotherapy on HRQOL before and after rehabilitation therapy.

Clarifying the characteristics of HRQOL, the relationship between HRQOL and ADL, and the factors that affect HRQOL in patients with brain tumors at the time of hospital discharge will provide useful information for implementing rehabilitation therapy. Further, investigating the EQ-5D-5L index score of patients with brain tumors may provide evidence for the cost-effectiveness of rehabilitation treatment in the future. In the present report, we investigated the effects of brain tumor type, recurrence, and treatment on the EQ-5D-5L index score. In addition, this study aimed to clarify the characteristics of HRQOL in different brain tumor types and its relationship with ADL.

## Methods

### Study design

This study uses a single-center cross-sectional study design. The design followed the international recommendations for Strengthening the Reporting of Observational Studies in Epidemiology [21]. This study was approved by the Niigata University Ethical Review Committee (Approval No.: 2020-0380). The authors obtained written informed consent from patients who were hospitalized between April and September 2021 and used an opt-out for subjects admitted between April 2016 and March 2021.

### Patients

The participants comprised patients aged ≥20 years who were admitted to The Niigata University Medical & Dental Hospital for the treatment of brain tumors between April 2016 and September 2021. The patients were also undergoing physical and occupational therapy, or physical and occupational therapy with speech therapy. The exclusion criteria were based on previous studies [19,22] and included: those who scored <23 on the Mini Mental State Examination, those who had difficulty answering the HRQOL questions due to aphasia or severe higher brain dysfunction, and those who had difficulty answering the questions due to poor general health.

### Assessment of general health and HRQOL

The FIM was used to assess ADL, and the Karnofsky performance status (KPS) was used to assess general health. HRQOL was assessed using the QLQ-C30, BN20, and EQ-5D-5L. All parameters were assessed at the time of discharge. FIM and KPS were assessed by the therapist in charge, whereas in principle, patients were required to answer the HRQOL questions by themselves. The therapist in charge of the patient was allowed to assist the patient in answering the questions if the patient had certain limitations that impaired them from filling out the form. These limitations included reading impairment due to motor paralysis or visual field impairment caused by central nervous system disorders. Age, sex, brain tumor type, tumor location (right, left, other), surgery, radiotherapy, chemotherapy, first occurrence or recurrence, and destination were extracted from the medical records. Brain tumors were classified based on the results of pathological examination according to the WHO 2016 Pathological Classification of Brain Tumors [23]. Finally, based on previous studies [15,22], the patients were classified into five groups: glioblastoma, grade III brain tumors (WHO grade III), primary central nervous system lymphomas (PCNSLs), MBTs, and grade I brain tumors (WHO grade I). The criteria used to determine whether surgery was performed were based on a previous study [24] and excluded biopsy from being considered a surgery. In order to examine the effect of treatment during hospitalization, we did not include a history of previous surgery, radiotherapy, or chemotherapy in patients with recurrence.

### Measurements

#### FIM

The FIM is an assessment of ADL, consisting of a motor category for self-care tasks (eating, grooming, bathing, dressing, toileting), sphincter control tasks (bladder management, bowel management), transfer tasks (bed-to-chair transfer, toilet transfer, tub or shower transfer), and locomotion tasks (walk or wheelchair, stairs), and a cognitive category for communication tasks (comprehension, expression) and social cognition tasks (social interaction, problem solving, memory). Each task is scored on a scale of 1 to 7 according to the level of independence, with 1 representing complete assistance and 7 representing complete independence. The total score ranges from 18 to 126, with a higher score indicating a greater degree of independence.

### QLQ-C30 and BN20

In this study, QLQ-C30 Japanese version (3rd edition) and BN20 were used for evaluation. These are the HRQOL questionnaires developed by the EORTC, which have been reported to be valid and reliable [4,5]. The QLQ-C30 is a disease-specific HRQOL assessment scale for patients with cancer. It consists of five functional scales (physical, role, cognitive, emotional functioning, and social functioning), nine symptom scales (nausea and vomiting, fatigue, dyspnea, pain, insomnia, appetite loss, constipation, diarrhea, and financial difficulties), and global health status.

The BN20 is a disease-specific measure of brain tumor symptoms. The BN20 is divided into the following symptom scales: future uncertainty, visual disorder, motor dysfunction, communication deficit, headache, seizure, drowsiness, hair loss, itchy skin, weakness of legs, and loss of bladder control.

The use of the above rating scales was approved by the EORTC Quality of Life Group. The QLQ-C30 and BN20 subscales are scored from 0 to 100 according to the scoring manual. A higher score on the QLQ-C30 functional scale and general health indicates better health, while a lower score on the QLQ-C30 symptom scale and BN20 indicates fewer complaints or better health.

#### EQ-5D-5L

The EQ-5D-5L is a generic preference-based measure of HRQOL developed by the EuroQol Group. EQ-5D-5L consists of five dimensions related to mobility, self-care, common activities, pain/discomfort, and anxiety/depression. Patients answer each item on a scale of 1 to 5 (no problems, slight problems, moderate problems, severe problems, and extreme problems). Initially developed by the EuroQol Group in 1987, the EuroQol-5Dimension-3Level (EQ-5D-3L) index was a five-item, three-level instrument. However, its sensitivity was insufficient, and a ceiling effect was identified. As a result, the five-level EQ-5D-5L was released to overcome these shortcomings [25]. In Japan, the EQ-5D-5L conversion table was completed in 2015, and the EQ-5D-5L index score reflecting Japanese values can be calculated [6]. The EQ -5D-5L utility index ranges from -0.025 to 1.00 (full health status).

### Statistical analysis

A one-way analysis of variance was performed to compare FIM total score, KPS, and HRQOL among the glioblastoma, WHO grade III, PCNSL, MBT, and WHO grade I groups. An unpaired t-test was used to compare the total FIM scores, KPS, and HRQOL between the two groups of patients who did and did not undergo surgery, radiotherapy, or chemotherapy, and those with first or recurrent disease. Pearson’s correlation analysis was used to investigate the relationship between the EQ-5D-5L index score, FIM, and disease-specific HQOL scale. In accordance with Guilford’s Rule of Thumb [26], the criterion for the strength of correlation was set as follows: |r| = 0-0.2 as “almost no correlation”, 0.2-0.4 as “weak correlation”, 0.4-0.7 as “moderately correlated”, and 0.7–1.0 as “strongly correlated”. Finally, a multiple regression analysis was performed to investigate the factors affecting the EQ-5D-5L index score at the time of hospital discharge, with the EQ-5D-5L index score as the dependent variable and age, sex, brain tumor type, surgery, radiotherapy, chemotherapy, and newly diagnosed or recurrent disease as independent variables. In this study, the forced imputation method of analysis was used to visually compare all independent variables with one other. The independent variables were selected with reference to previous studies that used the HRQOL scale and FIM total score as independent variables [15,27]. Categorical data were transformed into dummy variables, and WHO grade I was used as the reference category for brain tumor type. A P-value of <0.05 was regarded as being statistically significant, and all reported P-values were two-tailed. All statistical procedures were conducted using SPSS for Windows version 24.

## Results

### Patient characteristics

The patient characteristics are summarized in Table 1. The mean age of the patients was 61.1 years, and 59% were male. In addition, 74% of all patients were newly diagnosed, and 79% were discharged to home. The number of patients in each group was 21 in the glioblastoma group (27.6%), 10 in the WHO grade III group (13.2%), 10 in the PCNSL group (13.2%), 9 in the MBT group (11.8%), and 26 in the WHO grade I group (34.2%). None of the 10 patients in the PCNSL group had undergone surgical resection, whereas all patients in the MBT and WHO grade I groups had undergone surgery. In addition, none of patients in the WHO Grade I group received radiotherapy or chemotherapy.

**Table 1.**
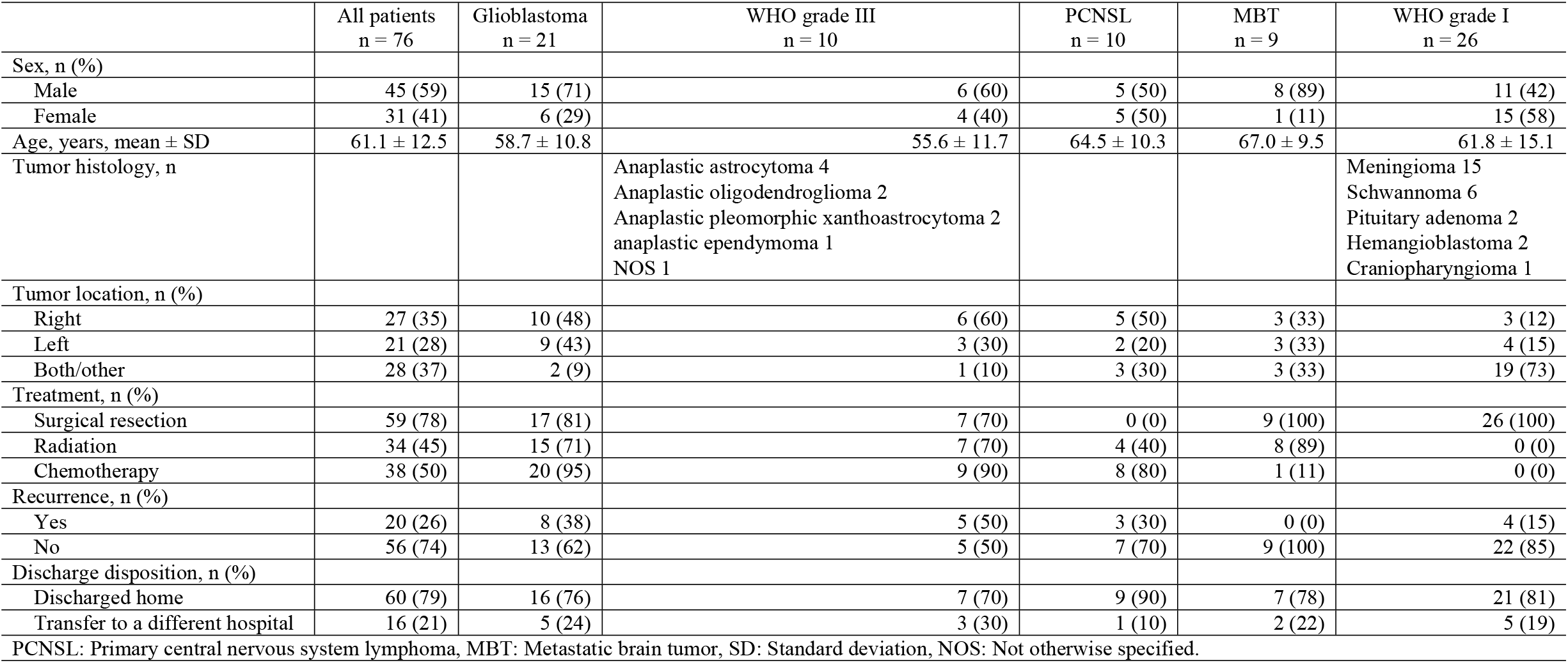
Patient characteristics.

### Comparison of assessment scores among brain tumor types

The assessment scores are summarized in Table 2. The FIM total score and KPS did not differ significantly among the brain tumor types. In contrast, the EQ-5D-5L index score (p = 0.048), emotional functioning (p = 0.015), financial difficulties (p = 0.002), and future uncertainty (p = 0.014) significantly differed among the groups. The EQ-5D-5L index score for all patients was 0.689±0.205. The glioblastoma group received the lowest score (0.574±0.229) and the WHO grade I group received the highest score (0.762±0.135). In addition, the glioblastoma group received the lowest score for emotional functioning and the highest scores for financial difficulties and future uncertainty among all groups.

**Table 2.**
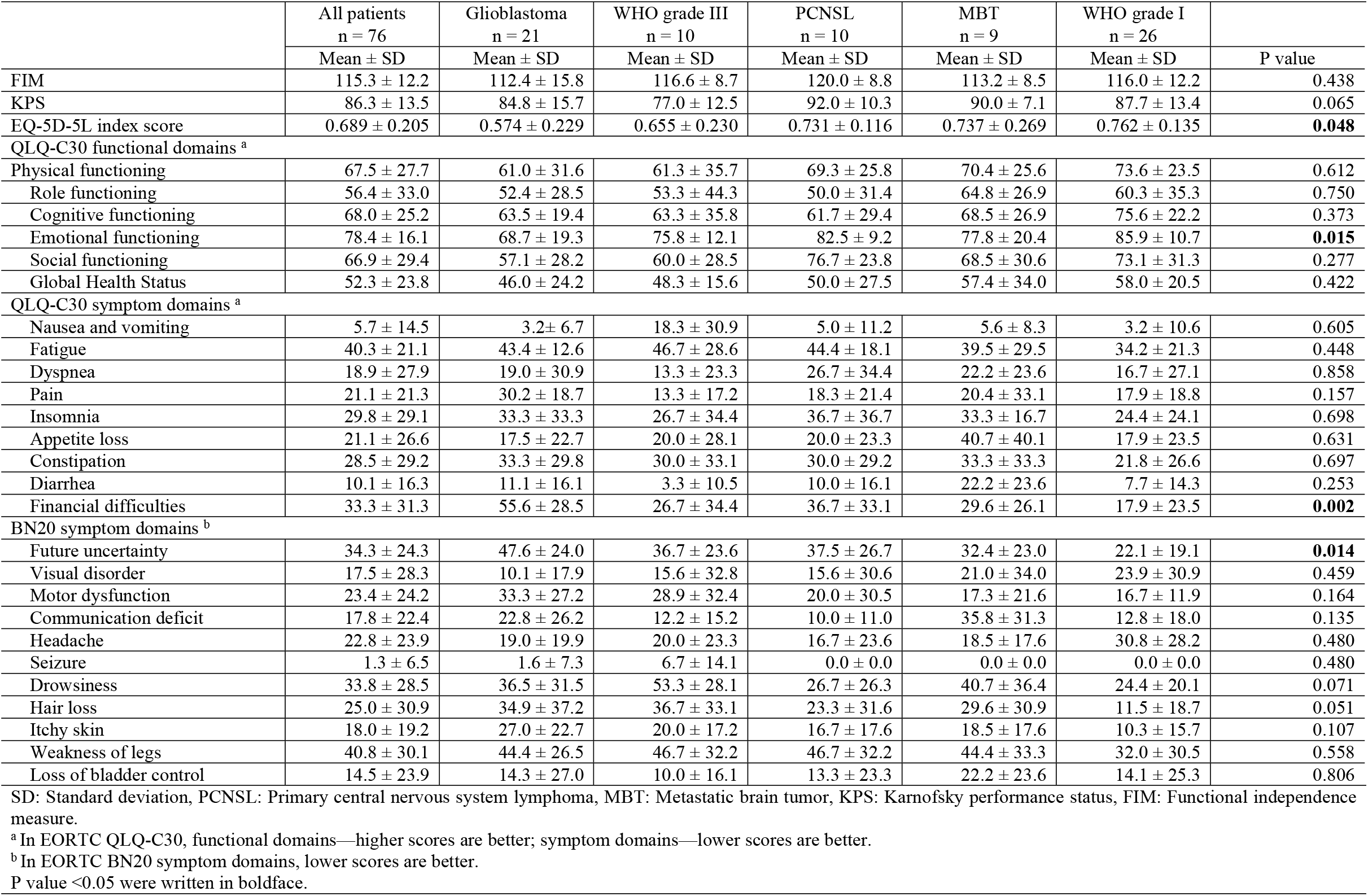
Comparison of FIM and KPS with HRQOL in tumor classification.

### Comparison of the different treatment groups and relapse and newly diagnosed groups

The results of the comparisons of the different treatment groups and relapse and newly diagnosed groups are presented in Table 3. The role functioning score was significantly higher (p = 0.027) and the scores for fatigue (p = 0.030), future uncertainty (p = 0.025), and weakness of legs (p = 0.020) were significantly lower in the group that underwent surgery than in the group that did not undergo surgery. The score for headache (p = 0.006) was significantly lower and the scores for hair loss (p = 0.001) and itchy skin (p = 0.002) were significantly higher in the group that received radiotherapy than in the group that did not receive radiotherapy. The EQ-5D-5L index (p = 0.029), emotional functioning (p = 0.027), and visual disorder (p = 0.038) scores were significantly lower and the financial difficulties (p = 0.013), future uncertainty (p = 0.044), hair loss (p = 0.018), and itchy skin (p = 0.002) scores were significantly higher in the group that received chemotherapy than in the group that did not receive chemotherapy. The KPS (p = 0.009), FIM total score (p = 0.048), EQ-5D-5L index score (p = 0.016), physical functioning score (p = 0.004), and role functioning score (p = 0.032) were significantly lower and the fatigue (p = 0.002), future uncertainty (p = 0.032), and drowsiness (p = 0.033) were significantly higher in the recurrence group than in the newly-diagnosed group.

**Table 3.**
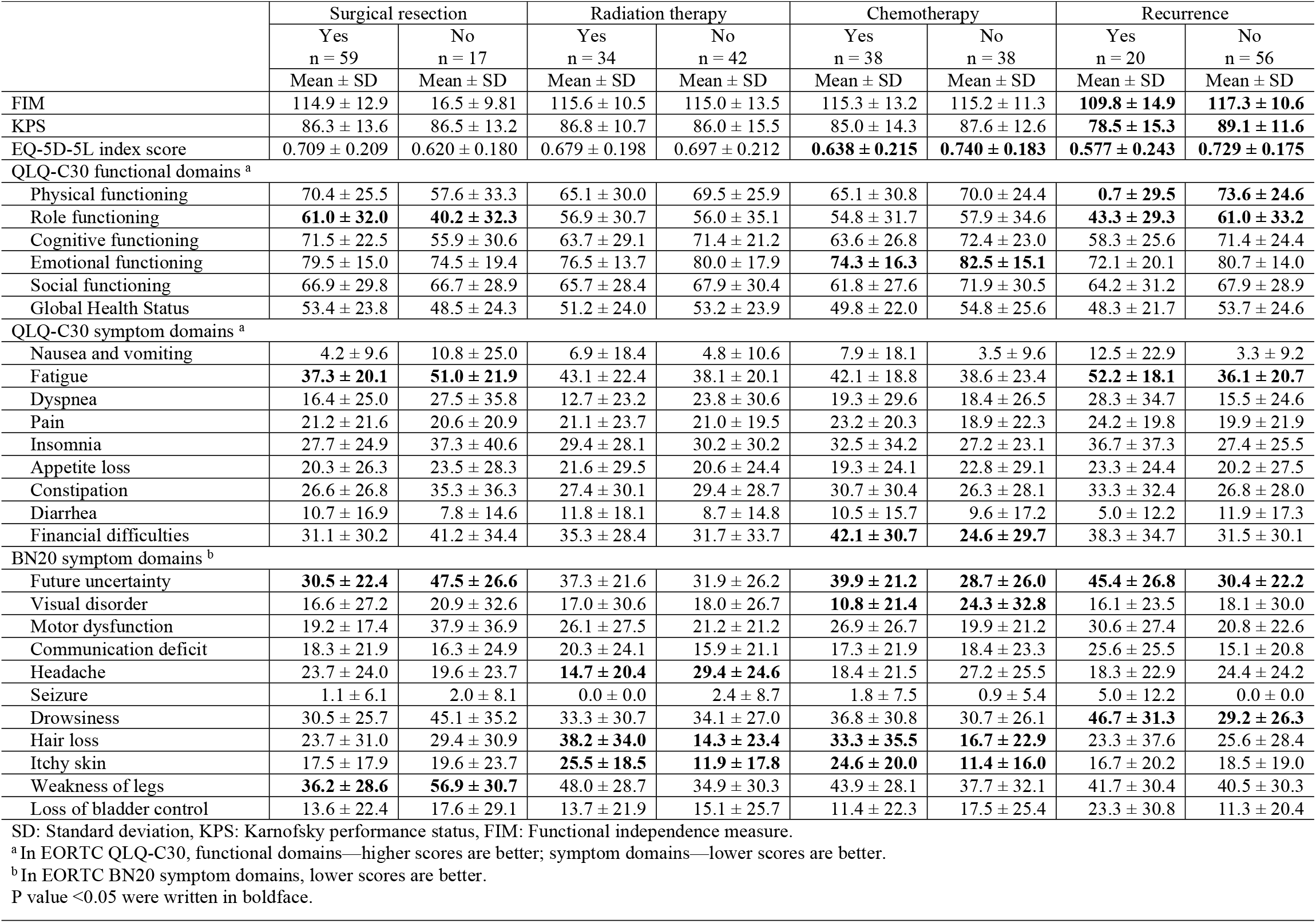
Comparison of FIM, KPS and HRQOL with and without each treatment and with and without recurrence.

### Correlations among the EQ-5D-5L index score, FIM, and disease-specific HRQOL scale

The correlations among the EQ-5D-5L index score, FIM, and disease-specific HRQOL scale are shown in Table 4. There was a strong correlation between the EQ-5D-5L index score and FIM (r = 0.627, p<0.001). Furthermore, the EQ-5D-5L index score and the disease-specific HRQOL scale showed significant correlations for all items with the exception of headache, hair loss, and itchy skin. In particular, strong correlations were observed with physical functioning (r = 0.723, p<0.001). In contrast, only physical functioning (r = 0.610, p<0.001) and dyspnea (r = -0.433, p<0.001) showed more than a moderate correlation between FIM and the disease-specific HRQOL measure.

**Table 4.** Correlation between EQ-5D-5L index score, FIM, and disease-specific HQOL scale.

|  | FIM | EQ-5D-5L index score |
| --- | --- | --- |
|  | Pearson's r | Pearson's r |
| EQ-5D-5L index score | **0.627\*\*** |  |
| QLQ-C30 functional domains <sup>a</sup> |  |  |
| Physical functioning | **0.610\*\*** | **0.723\*\*** |
| Role functioning | 0.325** | **0.602\*\*** |
| Cognitive functioning | 0.354** | **0.584\*\*** |
| Emotional functioning | 0.213 | **0.651\*\*** |
| Social functioning | 0.171 | **0.482\*\*** |
| Global Health Status | 0.129 | **0.430\*\*** |
| QLQ-C30 symptom domains <sup>a</sup> |  |  |
| Nausea and vomiting | -0.098 | -0.228* |
| Fatigue | -0.331** | **-0.669\*\*** |
| Dyspnea | **-0.433\*\*** | **-0.496\*\*** |
| Pain | -0.281* | **-0.533\*\*** |
| Insomnia | -0.184 | **-0.400\*\*** |
| Appetite loss | -0.139 | **-0.412\*\*** |
| Constipation | −0.110 | −0.274* |
| Diarrhea | −0.157 | −0.249* |
| Financial difficulties | −0.077 | **−0.406\*\*** |
| BN20 symptom domains <sup>b</sup> |  |  |
| Future uncertainty | −0.174 | **−0.566\*\*** |
| Visual disorder | −0.142 | −0.256* |
| Motor dysfunction | −0.095 | **−0.448\*\*** |
| Communication deficit | −0.145 | −0.374** |
| Headache | 0.115 | 0.003 |
| Seizure | 0.073 | −0.231* |
| Drowsiness | −0.180 | **−0.524\*\*** |
| Hair loss | 0.124 | −0.141 |
| Itchy skin | 0.081 | −0.030 |
| Weakness of legs | 0.017 | −0.273* |
| Loss of bladder control | −0.265* | −0.377** |

FIM: Functional independence measure.
<sup>a</sup> In EORTC QLQ-C30, functional domains—higher scores are better; symptom domains—lower scores are better.
<sup>b</sup> In EORTC BN20 symptom domains, lower scores are better.
Pearson's $r \geq |0.400|$ were written in boldface.
\*\* $p < 0.01$ , \* $p < 0.05$ .

### Multiple regression analysis for 5EQ-5D-5L index score

Multiple regression analysis was performed on the 5EQ-5D-5L index score, with age, sex, brain tumor type, surgery, radiotherapy, chemotherapy, and first occurrence or recurrence as independent variables (Table 5). Glioblastoma (standard partial regression coefficient: -0.570, p = 0.024) and surgery (standard partial regression coefficient: 0.376, p = 0.030) were identified as factors affecting the EQ-5D-5L index score.

**Table 5.** Multiple regression analysis with EQ-5D-5L index score as the dependent variable.

| | B | $\beta$ | 95% Confidence interval | | P value |
| --- | --- | --- | --- | --- | --- |
|  |  |  | Lower | Upper |  |
| Intercept | 0.684 |  | 0.341 | 1.027 | <0.001** |
| Age | −0.002 | −0.109 | −0.005 | 0.002 | 0.339 |
| Sex |  |  |  |  |  |
| Female (ref) |  |  |  |  |  |
| Male | −0.012 | −0.028 | −0.083 | 0.106 | 0.807 |
| Tumor histology |  |  |  |  |  |
| WHO grade I (ref) |  |  |  |  |  |
| Glioblastoma | -0.259 | -0.570 | -0.484 | -0.035 | 0.024* |
| WHO grade III | -0.151 | -0.250 | -0.388 | 0.086 | 0.209 |
| PCNSL | -0.087 | -0.144 | -0.165 | 0.339 | 0.495 |
| MBT | -0.115 | -0.183 | -0.296 | 0.065 | 0.201 |
| Surgical resection |  |  |  |  |  |
| No (ref) |  |  |  |  |  |
| Yes | 0.183 | 0.376 | 0.018 | 0.348 | 0.030* |
| Radiation therapy |  |  |  |  |  |
| No (ref) |  |  |  |  |  |
| Yes | 0.097 | 0.236 | -0.033 | 0.227 | 0.142 |
| Chemotherapy |  |  |  |  |  |
| No (ref) |  |  |  |  |  |
| Yes | 0.057 | 0.139 | -0.124 | 0.238 | 0.534 |
| Recurrence |  |  |  |  |  |
| No (ref) |  |  |  |  |  |
| Yes | -0.085 | -0.183 | -0.194 | 0.025 | 0.126 |

B: Partial regression coefficient, $\beta$ : Standardized regression coefficient, PCNSL: Primary central nervous system lymphoma, MBT: Metastatic brain tumor.
Adjusted R<sup>2</sup>: 0.188.
\*\*p < 0.01, \*p < 0.05.

## Discussion

This study aimed to investigate the effects of brain tumor type, recurrence, and treatment on the EQ-5D-5L index score and to clarify the characteristics of HRQOL in different brain tumor types and its relationship with ADL.

The mean EQ-5D-5L index score at the time of hospital discharge for all patients with brain tumors in this study was 0.689±0.205 (mean age 61.1 years). The WHO grade I group had the highest score of 0.762±0.135 (mean age 61.8 years) and the glioblastoma group had the lowest score of 0.574±0.229 (mean age 58.7 years). In Japan, the EQ-5D-5L index score for patients with brain tumors has not been reported previously, and in other countries, Wagner et al [7] reported a mean index score of 0.72 in 3-month postoperative patients with benign meningiomas. The EQ-5D-5L index score of the WHO grade I group in this study was comparable, although simple comparison is difficult because the EQ-5D-5L index score is calculated using a country-specific conversion table. However, the mean EQ-5D-5L index score of the general population in Japan was reported to be 0.936 in the 50s and 0.911 in the 60s [28]. In the case of patients with various types of outpatient cancers aside from brain tumors, the reported value was 0.827 [29]. In addition, the mean score was 0.52 (mean age 57 years) in stroke patients, who are expected to present with similar functional impairment [30]. The EQ-5D-5L index score of the brain tumor patients in this study was lower than that of the general population and patients with other cancers, although the results should be interpreted with caution regarding the different effects of the time of assessment, age, and disease. Furthermore, the values were similar between the current glioblastoma group and previous reports of stroke. In the present study, there were significant differences in emotional functioning, financial difficulties, and future uncertainty among brain tumor types. In addition, the glioblastoma group showed the lowest values for all scales. Budrukkar et al [22] reported that the Global Health Status of the QLQ-C30 was significantly lower in the high-grade glioma (HGG) group than in the low-grade glioma (LGG) group. In a study of glioma patients treated with rehabilitation therapy during hospitalization, Umezaki et al [27] found that the HGG group had fewer complaints of QLQ-C30 constipation and more complaints of BN-20 hair loss and itchy skin than did the LGG group. These previous studies and the current results differed in the items that showed significant differences. This may have been due to the differences in the brain tumor type and the individuality of the hospitals in the study area. However, it is interesting to note that in the present study, the scores of the HRQOL items reflecting psychological aspects were lower in the glioblastoma group than in the other groups. Glioblastomas carry a poorer prognosis than other brain tumors, aside from MBTs, and may cause psychological problems. Indeed, patients with glioblastoma have been reported to have more depressive symptoms than patients with gastric, urological, breast, and lung cancers [31]. Further, most brain tumors classified as WHO grade I can be treated with surgery alone, but brain tumors classified as WHO grade II or higher often require radiation therapy or chemotherapy in addition to surgery. Moreover, these treatments may be continued after hospital discharge. These factors may be related to the emotional functioning, financial difficulties, and future uncertainty scores of the glioblastoma group. However, within the scope of this study, we have not been able to examine the above points, and they are only inferred.

A previous study in acute stroke patients reported a significant correlation between the EQ-5D-5L index score and FIM motor items [32] and Barthel index [33]. In contrast, a previous study of patients with brain tumors reported no correlation between the total FIM score at discharge and the Functional Assessment of Cancer Therapy-Brain (FACT-Br), a disease-specific HRQOL scale [18]. In the present study, we also found a significant correlation between the FIM and EQ-5D-5L index score. However, in the FIM total score and disease-specific HRQOL scale, the items that showed significant correlations were limited to those related to physical function. This finding was similar to those of previous studies, although the HRQOL scale used was different. However, our results are noteworthy in that the EQ-5D-5L index score and the disease-specific HRQOL scale showed significant correlations for all items with the exception of headache, hair loss, and itchy skin on the BN20. Hirose et al [29] reported a correlation between changes in adverse events and EQ-5D-5L index scores in patients with cancer. In addition, the EQ-5D-3L index score of patients with brain tumors is reportedly associated with the emotional well-being item of the FACT-Br [34] and anxiety and depression symptoms [35]. The correlations between EQ-5D-5L index scores and the QLQ-C30 and BN20 in this study were similar to those in previous studies, although the target diseases and HRQOL assessment scales were different. Coomans et al [24] reported the impact of HRQOL on OS in patients with gliomas, but the added value was low, indicating the limitations of using HRQOL as a prognostic indicator of OS. However, Edelstein et al [31] stated that the limitation of activity and participation due to glioblastoma is a factor that interferes with subjective well-being and mentioned the possibility of rehabilitation therapy to improve HRQOL. Similarly, in addition to training to improve ADL as indicated by the HRQOL assessment, the importance of rehabilitation treatment for patients with brain tumors, which is largely affected by individual complaints, is demonstrated in this study.

Furthermore, we investigated the influence of factors such as brain tumor type, surgery, radiotherapy, chemotherapy, and recurrence on the EQ-5D-5L index score. The results of multiple regression analysis showed that glioblastoma and surgery were the most influential factors. It has been reported that surgery in patients with brain tumors prolongs survival and improves EQ-5D-3L index scores more in patients who undergo tumorectomy than in those who undergo biopsy [36,37]. Furthermore, tumorectomy has been suggested to improve HRQOL by providing a mass effect and improvement in hydrocephalus [22]. These previous findings support the results of the present study that surgery was a factor in improving the EQ-5D-5L index score. Vera et al [38] investigated the effect of different brain tumor classifications on the EQ-5D-3L index score in patients with gliomas who were undergoing outpatient treatment. After dividing the patients into two groups, grade II/III and grade IV, we reported that the grade of the brain tumor was not a factor affecting the EQ-5D-3L index score. Similar results were also reported in a study of postoperative HGG and LGG patients [39]. In the present study, glioblastoma reduced the EQ-5D-5L index score, and this finding differed from those of previous studies. However, these previous studies were limited to the glioma population. In our study, we used the EQ-5D-5L index and further divided the brain tumor classifications into five groups, which we believe is a new finding.

## Limitations

There are several limitations to this study. First, this study was conducted at a single institution and was limited to patients with brain tumors who underwent rehabilitation treatment. In addition, patients with poor general health, cognitive decline, or aphasia were excluded. Therefore, the results are not generalizable to all patients with brain tumors. In addition, because this was a cross-sectional study, we were not able to compare the findings before and after rehabilitation treatment, nor were we able to examine changes after hospital discharge.

Second, brain tumor type may affect the choice of treatment for newly diagnosed and recurrent brain tumors. In the PCNSL group, the main treatment was chemotherapy and radiation therapy without radical surgical resection. For patients with other tumor types, it is important to removing as much tumor as possible, but complete tumor resection cannot be performed if the tumor is located in the brainstem or similar areas, or if the disease is intractable, including distant recurrence. Thus, the group of patients who did not undergo surgery may have included more difficult-to-treat cases, which may have influenced the results. Third, previous studies have reported that a history of epilepsy and impaired cognitive function [40], as well as tumor location and size [41], affect HRQOL, but we were unable to examine their effects in this study. In addition, the late effects of radiotherapy and chemotherapy may have affected HRQOL after the study. Since brain tumors are rare, there is a need to evaluate a greater number of cases by conducting multicenter studies. Furthermore, in the present study, the analysis was only performed at discharge after rehabilitation therapy. We are currently conducting continuous surveys before and after rehabilitation treatment and after discharge from the hospital in order to longitudinally understand the effect of rehabilitation on ADL and HRQOL.

## Conclusion

This study investigated the HRQOL of patients with brain tumors who underwent rehabilitation therapy and investigated the factors affecting the EQ-5D-5L index score from various perspectives, including various brain tumor types, treatment methods, and recurrence. In addition, we examined the relationship between the EQ-5D-5L index score, disease-specific HRQOL scale, and FIM total score. The EQ-5D-5L index score of the patients in this study was lower than that of the general adult population. In addition, the glioblastoma group had the lowest EQ-5D-5L index score among all brain tumor types. In addition, the EQ-5D-5L index score was significantly correlated with most of the items of the disease-specific HRQOL scale in addition to the total FIM score. Multiple regression analysis revealed that glioblastoma and surgery were factors that significantly influenced the EQ-5D-5L index score. The results of our study may provide useful information for the rehabilitation of patients with brain tumors.

## Data Availability

Data cannot be shared publicly because of privacy for the patients.

## Acknowledgments

We would like to express our sincere gratitude to the patients who cooperated in this study, and to the Department of Neurosurgery and Rehabilitation Medicine and therapists at The Niigata University Medical & Dental Hospital.

